# Predictors of Preeclampsia and Pregnancy Associated Outcomes Among Pregnant Women: A Prospective Cohort Study in Selected Hospitals in Kiambu County, Kenya

**DOI:** 10.64898/2026.10.06.26364837

**Authors:** Joseph Mukala, Dominic Mogere, Winnie Muniu, Beatrice Mwangi, Esther Wanjira, Robin Nyakundi, Julius Wairimu

## Abstract

Preeclampsia is a multisystem hypertensive disorder of pregnancy that emerges after 20 weeks of gestation, and remains a leading cause of maternal and perinatal morbidity and mortality globally, yet evidence on its predictors and outcomes in Kenya remains limited. This prospective cohort study compared pregnant women with and without preeclampsia who attended selected hospitals in Kiambu County, to determine the predictors of preeclampsia and its associated maternal and neonatal outcomes. The results indicated a very strong statistical association between preeclampsia and the following i.e parity, time taken to reach the hospital, hypertension and diabetes mellitus (p-value <0.001) and religion (p-value <0.013). Women with preeclampsia had nearly twice the risk and six times the odds of delivering low-birth-weight infants compared with women without preeclampsia (RR = 1.97; OR = 6.25). A strong statistically significant association was observed between multiparity and prematurity (RR = 5.3, OR = 8.4, p < 0.001). Multiparous women with preeclampsia had approximately 5.3 times the risk and 8.4 times the odds of experiencing a premature birth compared with women without preeclampsia. In conclusion preeclampsia significantly increased adverse maternal and neonatal outcomes, highlighting the need for early high-risk pregnancies identification and strengthened antenatal care to improve both maternal and neonatal outcomes.

## Objectives

The study aimed at determining the predictors of preeclampsia among pregnant women attending selected hospitals in Kiambu County, Kenya. It further sought to compare maternal, neonatal, clinical, laboratory, and obstetric characteristics between women with and without preeclampsia.

## Background

Preeclampsia is one of the most serious hypertensive disorders of pregnancy and remains a major contributor to maternal and neonatal morbidity and mortality worldwide because of its complex pathophysiology, unpredictable progression, and association with severe multisystem complications [1,2]. Globally, hypertensive disorders complicate nearly one in ten pregnancies, while preeclampsia affects approximately 3–8% of all pregnancies and contributes substantially to maternal deaths, preterm birth, fetal growth restriction, low birth weight, and neonatal mortality, particularly in low-resource settings where access to quality obstetric care remains inadequate [2,3,4]. In sub-Saharan Africa, the burden continues to rise, with recent evidence identifying preeclampsia as a leading cause of preventable maternal and perinatal deaths despite advances in antenatal care.

Kenya continues to experience an unacceptably high maternal mortality ratio of approximately 355 deaths per 100,000 live births, with hypertensive disorders, particularly eclampsia and preeclampsia, ranking among the leading direct obstetric causes of maternal mortality [5,2]. National reports indicate that eclampsia contributes approximately 13–14% of maternal deaths, while facility-based studies at Kenyatta National Hospital have shown that hypertensive disorders account for more than half of direct maternal deaths [6,7,8]. Kiambu County, one of Kenya’s rapidly urbanizing counties with a high demand for maternal health services, continues to face challenges related to timely diagnosis, referral, and management of obstetric emergencies despite improvements in healthcare infrastructure [9]. Although several studies have described the epidemiology of preeclampsia in Kenya, prospective evidence examining its predictors together with pregnancy-associated maternal and neonatal outcomes remains scarce. This study therefore sought to determine the predictors and pregnancy associated outcomes of preeclampsia among pregnant women attending selected hospitals in Kiambu County to generate evidence that can strengthen antenatal risk stratification, early detection, and clinical management of high-risk pregnancies.

## Methods

Sample size was calculated using the two-independent-proportion method for a binary pregnancy outcome, assuming a two-sided 5% significance level, 95% confidence, 80% power, and equal cohort allocation. Previous estimates of low birth weight were 22.6% among pre-eclamptic women and 4.65% among uncomplicated pregnancies. After accounting for 15% attrition, recruitment was increased to 88 participants per cohort. This was a prospective cohort study conducted from April to December 2022 under License No. NACOSTI/P/22/16777. A total of 176 pregnant women were enrolled across all pregnancy trimesters. Participants were further subdivided into two groups, as whether they had preeclampsia meaning that their pressure value ≥ 140/90 mmHg or no. Thereafter, the first arm comprised of 88 pregnant women who had confirmed high blood pressure, and each case was allocated a control in the second arm, either 88 pregnant women with normal blood pressure range (<140/90). Both groups were systematically followed until delivery, with standardized data collection on sociodemographic, clinical characteristics, investigations, as well as maternal and neonatal outcomes. This methodological approach allowed for rigorous comparison between affected and unaffected pregnancies, thereby generating evidence on the burden and consequences of preeclampsia in a resource-constrained setting [10].

## Results

The study demonstrated a statistically significant association between preeclampsia and the following: parity (p = 0.001), religion (p = 0.013), the time taken to reach healthcare facilities (p < 0.001). Although maternal age was not statistically associated with preeclampsia, women aged ≥34 years had a relatively higher prevalence of the condition in the case group 30 (17.0%), compared to control group 15 (8.6%). Clinically, pervaginal bleeding (p = 0.007), lower limb edema, headache (p < 0.001), and blurred vision (p < 0.014). Ultrasound utilization (p < 0.039), the timing of ultrasound examination (p < 0.001). Laboratory findings; urinary leucocytes, haemoglobin levels and blood glucose status (p < 0.001). In contrast, proteinuria and HIV (p = 0.244, 0.497) respectively were not significantly associated with preeclampsia, although the interpretation of proteinuria findings should be made cautiously because over 40% of participants lacked documented proteinuria testing. In fact, preeclampsia was strongly associated with birth weight (p < 0.001), while the prevalence of low birth weight was estimated at 22%. Women with preeclampsia had nearly twice the risk of delivering low-birth-weight infants compared with women without preeclampsia (RR = 1.97; OR = 6.25). Cesarean section was more frequent among women with preeclampsia (25.1%) than among those without preeclampsia (21.1%), whereas spontaneous vaginal delivery was more common in the non-preeclampsia group (29.1% vs. 24.6%). APGAR score was significantly associated with preeclampsia (p < 0.011). Women with preeclampsia had nearly twice the risk and seven times odds of poor APGAR scores (RR = 1.85; OR = 7.38), sixfold higher risk of fetal distress (OR=6.25), preeclamptic multiparous women had approximately 5.3 times the risk and 8.4 times the odds of experiencing a premature birth compared with women without preeclampsia (RR = 5.3, OR = 8.4, p < 0.001). Equally, there was fourfold higher risk of neonatal death (OR: 4.26) compared with the non-preeclampsia group. Preeclampsia was associated with a more than two-weeks normal gestational reduction at delivery, with women in the preeclampsia group delivering from 30 weeks compared with an average of 39 weeks in the non-preeclampsia group.

## Discussion

The present study found a significant association between parity and preeclampsia (p = 0.001), with multigravida and grand multigravida women exhibiting a higher prevalence of the condition than women without preeclampsia, those multigravida and grand multigravida had also a very significant risk and odds of premature birth compared to women without preeclampsia. This finding suggests that higher parity is an important predictor of preeclampsia in the study setting. While preeclampsia has traditionally been regarded as a disorder primarily affecting nulliparous women because of immunological maladaptation during the first pregnancy [10], accumulating evidence indicates that grand multiparity has become an increasingly important risk factor, particularly in low- and middle-income countries. A systematic review by [11] reported that grand multiparity was associated with increased odds of preeclampsia in sub-Saharan Africa, largely due to advancing maternal age, chronic comorbidities, and cumulative vascular damage. Similar findings have been reported in Ethiopia [12], Nigeria [13], Tanzania [14], and Uganda [15]. The observed association may be explained by progressive endothelial dysfunction, chronic inflammation, oxidative stress, and impaired angiogenic balance resulting from repeated pregnancies. Alterations in angiogenic biomarkers, including placental growth factor (PlGF), soluble fms-like tyrosine kinase-1 (sFlt-1), and soluble endoglin, have been implicated in defective placentation and endothelial injury among women at increased risk of preeclampsia [16,17]. These findings suggest that the effect of parity on preeclampsia should be interpreted alongside maternal age, underlying chronic diseases, and cumulative cardiovascular risk rather than as an isolated obstetric characteristic.

Religion was significantly associated with preeclampsia (p = 0.013). However, this relationship is unlikely to reflect a direct biological effect but rather the influence of social and cultural determinants of health. Religious affiliation may shape healthcare-seeking behavior, antenatal care utilization, dietary practices, social support, and decision-making, which can indirectly affect pregnancy outcomes. Similar observations have been reported in Ghana [18], Nigeria [19], and Pakistan [20] while recent epidemiological evidence suggests that such associations are often attenuated after adjustment for socioeconomic and behavioral factors. Therefore, the observed association should be also interpreted cautiously, and further qualitative research is needed to clarify the underlying sociocultural mechanisms [21]. Further qualitative and mixed-method research studies are therefore needed to deepen the understanding between sociocultural pathways and religious affiliation.

Time taken to reach a health facility was significantly associated with preeclampsia (p < 0.001). This corroborated with previous findings reported by the [14,22], women living within one hour of a health facility were more frequently diagnosed with preeclampsia, likely reflecting earlier detection, referral patterns, and hospital-based selection bias rather than a true increase in disease risk. A recent WHO analysis involving 54 low-and-middle income countries reported that prolonged travel time significantly increased the risk of maternal complications due to delays in diagnosis and treatment [22]. Similarly, analyses from Ethiopia [23], Tanzania [14], Kenya Demographic and Health Survey [5], showed that women residing far from health facilities had poorer maternal outcomes. Therefore, the findings of the study underscore the importance of considering healthcare accessibility not merely as geographical distance but also as diagnostic opportunity and health system responsiveness.

Pre-existing hypertension was strongly associated with preeclampsia (p < 0.001), with nearly one in four women having a higher likelihood of developing the condition. This finding is consistent with previous evidence identifying chronic hypertension as a major predictor of preeclampsia [3,22,24,25]. However, some studies have reported lower progression rates among women receiving early antihypertensive treatment and intensive antenatal monitoring [26].

Diabetes mellitus was significantly associated with preeclampsia (p < 0.001), although 11.9% of women with pre-existing diabetes did not develop preeclampsia. This finding agrees with previous evidence identifying diabetes as a major risk factor for preeclampsia [22,27,28]. However, some diabetic women remain unaffected, possibly due to effective glycemic control, early management, or study-related methodological differences [29].

Maternal age was not significantly associated with preeclampsia (p = 0.087), although women aged ≥34 years had a relatively higher prevalence of the condition. This finding is consistent with evidence identifying advanced maternal age as a risk factor for preeclampsia [22,30,31]. However, the lack of statistical significance may reflect the study’s limited sample size and insufficient statistical power rather than the absence of an age-related effect. Pervaginal bleeding was significantly associated with preeclampsia (p<0.007), occurring exclusively among affected women. This finding is consistent with studies linking antepartum hemorrhage to severe preeclampsia and placental pathology [32,33]. However, lower rates have been reported where early antenatal surveillance and timely management prevented severe placental complications [34].

Lower limb edema showed the strongest association with preeclampsia (p < 0.001). This finding is consistent with studies reporting edema as a common clinical manifestation of severe preeclampsia [11,32]. However, its diagnostic value remains limited because peripheral edema frequently occurs in normal pregnancy [25,35].

Blurred vision was significantly associated with preeclampsia (p = 0.014). This finding agrees with studies identifying visual disturbances as an important manifestation of severe preeclampsia and disease progression [11,33]. However, lower frequencies have been reported where early antenatal detection prevented neurological complications [34].

Moreover, time of ultrasound utilization was significantly associated with preeclampsia (p<.001). This finding supports evidence that routine obstetric ultrasonography enhances early identification of high-risk pregnancies and improves obstetric surveillance [33,36,37]. However, in the current study women with preeclampsia underwent second trimester ultrasound either in the case group 34(19%) against 46(26%) in the control group, where approximately 35 (19.9%) reported that ultrasound was never done. None significance association was reported in settings with near-universal ultrasound coverage [38]. This finding supports evidence that early ultrasound and antenatal care improve risk stratification, and may facilitate timely prevention of preeclampsia [11,31,32]. In fact, studies from higher income countries suggested that ultrasound timing alone may have limited predictive value when comprehensive antenatal screening is universally implemented [31].

Interestingly, presence of leucocytes in the urine was significantly associated with preeclampsia (p < 0.001). This finding supports evidence linking urinary tract infections and renal inflammation with an increased risk of preeclampsia through inflammatory mechanisms [39,40]. However, other studies found no independent association after adjusting for confounding factors [41].

Haemoglobin levels were significantly associated with preeclampsia (p < 0.001). This finding is consistent with evidence describing hemoconcentration as a characteristic feature of preeclampsia resulting from plasma volume contraction [16]. Other studies have associated maternal anaemia with an increased risk of preeclampsia [42].

Blood glucose test was significantly associated with preeclampsia (p < 0.001). This finding is consistent with evidence linking impaired glucose metabolism to an increased risk of hypertensive disorders during pregnancy [43,44,29,45]. However, some studies have reported weaker associations after adjustment for maternal obesity and other metabolic risk factors [44].

No significant association was observed between HIV infection and preeclampsia (p = 0.497), and no association could be established for VDRL because all participants tested negative. This finding agrees with studies reporting no independent relationship between HIV and preeclampsia after adjustment for antiretroviral therapy and indicating no direct causal link between syphilis and preeclampsia [36,46]. However, other studies have reported increased preeclampsia risk among women receiving highly active antiretroviral therapy [47,48].

Low birth weight was significantly associated with preeclampsia (p < 0.001), with affected women having nearly twice the risk (RR = 1.97) and sixfold higher odds (OR = 6.25) of delivering low-birth-weight infants. This finding is consistent with recent evidence attributing fetal growth restriction to placental insufficiency and uteroplacental hypoperfusion in preeclampsia [22,25,49,51]. Studies from settings with early aspirin prophylaxis and intensive fetal surveillance have reported substantially lower rates of low birth weight among women with preeclampsia [26,31].

Cesarean section was more common among women with preeclampsia, whereas spontaneous vaginal delivery predominated among normotensive women, despite lack of statistical significance. This finding agrees with reports that preeclampsia frequently necessitates operative delivery because of maternal or fetal compromise [22,25,31,33,45]. However, other studies have demonstrated that carefully selected women with stable preeclampsia can safely achieve vaginal delivery under appropriate obstetric management [52,53].

APGAR score was significantly associated with preeclampsia (p = 0.011), with affected women experiencing nearly twice the risk (RR = 1.85) and sevenfold higher odds (OR = 7.38) of fetal distress. This finding supports evidence linking placental dysfunction to neonatal compromise at birth [22,25,32,54]. However, improved intrapartum monitoring and timely neonatal resuscitation have been associated with better APGAR outcomes despite maternal preeclampsia [26,52]. Prematurity occurred four times more frequently among multiparous women with preeclampsia than among those without the condition. This finding concurs with evidence showing that medically indicated early delivery is often required to prevent severe maternal and neonatal complications [36,25,31,33,55]. However, improved expectant management in specialized obstetric units has reduced preterm delivery without increasing adverse maternal outcomes in selected women [52,53].

Perinatal death was four times more frequent among women with preeclampsia than among women without the condition. This finding agrees with evidence that severe placental dysfunction and prematurity substantially increase neonatal mortality in hypertensive pregnancies [22,32,45,54]. However, studies from well-resourced neonatal intensive care settings have reported significantly lower neonatal mortality through timely obstetric intervention and advanced neonatal care [26,52].

## Conclusion

This study demonstrated that preeclampsia was significantly associated with several sociodemographic, clinical, laboratory, and obstetric factors, including parity, religion, healthcare accessibility, pre-existing hypertension, diabetes mellitus, lower limb edema, headache, blurred vision, time of ultrasound utilization, urinary leucocytes, haemoglobin levels, and blood glucose status. Compared with women without preeclampsia, affected women experienced substantially poorer maternal and perinatal outcomes, including increased cesarean section delivery, low birth weight, fetal distress, poor APGAR scores, prematurity, and neonatal death, underscoring the considerable burden of preeclampsia on maternal and perinatal health. Therefore, preeclampsia remains a major contributor to adverse maternal and neonatal outcomes and is strongly associated with identifiable maternal risk factors and clinical indicators. Early risk stratification, timely diagnosis, and evidence-based obstetric management are essential to improving pregnancy outcomes.

Table 1 above showed that majority of the respondents were in their third trimester, aged between 26-33 years primigravida and took less than one hour to reach the facility.

**Table 1.** Socio-demographic Characteristic Associated with Preeclampsia

| **Variables** | **Overall: N =**<br>**176, %** | **Preeclampsia n = 88,**<br>**%** | **Non-Preeclampsia: n =**<br>**88, %** | **p-value** |
| --- | --- | --- | --- | --- |
| **Gestational Age** |  |  |  | 0.119 |
| first trimester 1-12 weeks | 5 (2.8) | 4 (2.3) | 1 (0.6) |  |
| second trimester 13-26 weeks | 42 (23.9) | 25 (14.2) | 17 (9.7) |  |
| third trimester 27-40 weeks | 129 (73.3) | 59 (33.5) | 70 (39.8) |  |
| **Age of the Respondent** |  |  |  | 0.087 |
| Less than 18 | 2 (1.1) | 1 (0.6) | 1 (0.6) |  |
| 18-25 | 50 (28.4) | 19 (10.8) | 31 (17.6) |  |
| 26-33 | 79 (44.9) | 38 (21.6) | 41 (23.3) |  |
| 34-41 | 41 (23.3) | 27 (15.3) | 14 (8.0) |  |
| 42-49 | 4 (2.3) | 3 (1.7) | 1 (0.6) |  |
| **Parity of the Respondent** |  |  |  | 0.001 |
| Primigravida | 77 (43.8) | 27 (15.3) | 50 (28.4) |  |
| Multigravida | 99 (56.3) | 61 (34.7) | 38 (21.6) |  |
| **Religion** |  |  |  | 0.013 |
| Catholic | 50 (28.4) | 32 (18.2) | 18 (10.2) |  |
| Protestant | 116 (65.9) | 55 (31.3) | 61 (34.7) |  |
| Muslim | 8 (4.5) | 1 (0.6) | 7 (4.0) |  |
| Hindu | 2 (1.1) | 0 (0.0) | 2 (1.1) |  |
| **Time Taken to Reach the Health Facility** |  |  |  | <.001 |
| less than 1 hour | 101 (57.4) | 59 (33.5) | 42 (23.9) |  |
| 1-2 hours | 45 (25.6) | 26 (14.8) | 19 (10.8) |  |
| 2-3 hours | 30 (17.0) | 3 (1.7) | 27 (15.3) |  |

Table 2 above showed that among non-preeclamptic women, 21 (12%) had a history of pre-existing diabetes mellitus, while 37 (21%) had a history of pre-existing hypertension. Diabetes mellitus and hypertension had a p<0.001.

**Table 2.** Historical Records of Chronic Diseases Associated with Preeclampsia

| Variables | Overall, N =<br>176 | Preeclampsia, N =<br>88 | Non-Preeclampsia, N<br>= 88 | p-value |
| --- | --- | --- | --- | --- |
| **Diabetes Mellitus** |  |  |  | **<.001** |
| Yes | 21 (11.9) | 0 (0.0) | 21 (11.9) |  |
| No | 155 (88.1) | 88 (50.0) | 67 (38.1) |  |
| **Hypertension** |  |  |  | **<.001** |
| Yes | 37 (21.0) | 34 (19.3) | 3 (1.7) |  |
| No | 139 (79.0) | 54 (30.7) | 85 (48.3) |  |

Table 3 above showed that among pregnant women with preeclampsia headache and lower limb edema were the commonest symptoms with 53(30.3%) against 52(29.5%), while blurred vision and pervaginal bleeding were less common 8(4.65); 24(13.7%) respectively. Pervaginal bleeding (p <0.007), Lower Limb edema and headache (p<0.001), Blurred Vision (p<0.014)

**Table 3:** Clinical Manifestation Associated with Preeclampsia

| Variables | Overall, N<br>= 176 | Preeclampsia, N<br>= 88 | Non-Preeclampsia, N<br>= 88 | p-value |
| --- | --- | --- | --- | --- |
| Pervaginal bleeding |  |  |  | 0.007 |
| Yes | 8 (4.5) | 8 (4.6) | 0 (0.0) |  |
| No | 168 (95.5) | 80 (45.5) | 88 (50.0) |  |
| Lower Limb edema |  |  |  | <.001 |
| Yes | 52 (29.5) | 43 (24.4) | 9 (5.1) |  |
| No | 124 (70.5) | 45 (25.6) | 79 (44.9) |  |
| Headache |  |  |  | <.001 |
| Yes | 53 (30.3) | 38 (21.6) | 15 (8.5) |  |
| No | 123 (69.9) | 50 (28.4) | 73 (41.5) |  |
| Epigastric Pain |  |  |  | 1.000 |
| Yes | 34 (19.3) | 17 (9.7) | 17 (9.7) |  |
| No | 142 (80.7) | 71 (40.3) | 71 (40.3) |  |
| Blurred Vision |  |  |  | 0.014 |
| Yes | 24 (13.7) | 18 (10.2) | 6 (3.4) |  |
| No | 152 (86.4) | 90 (39.8) | 82 (46.6) |  |

Table 4 above showed that most ultrasound examinations occurred during the second trimester. Prediabetes and anaemia were more frequent among women with preeclampsia 46(26.1%) against 34(19%), majority of prediabetic pregnant women were 27(15.3%) in preeclampsia, timing of ultrasound, leucocyturia, urinary pus, blood glucose, and full haemogram were significantly associated with preeclampsia (p<0.001), although several tests remained underutilized.

**Table 4:**
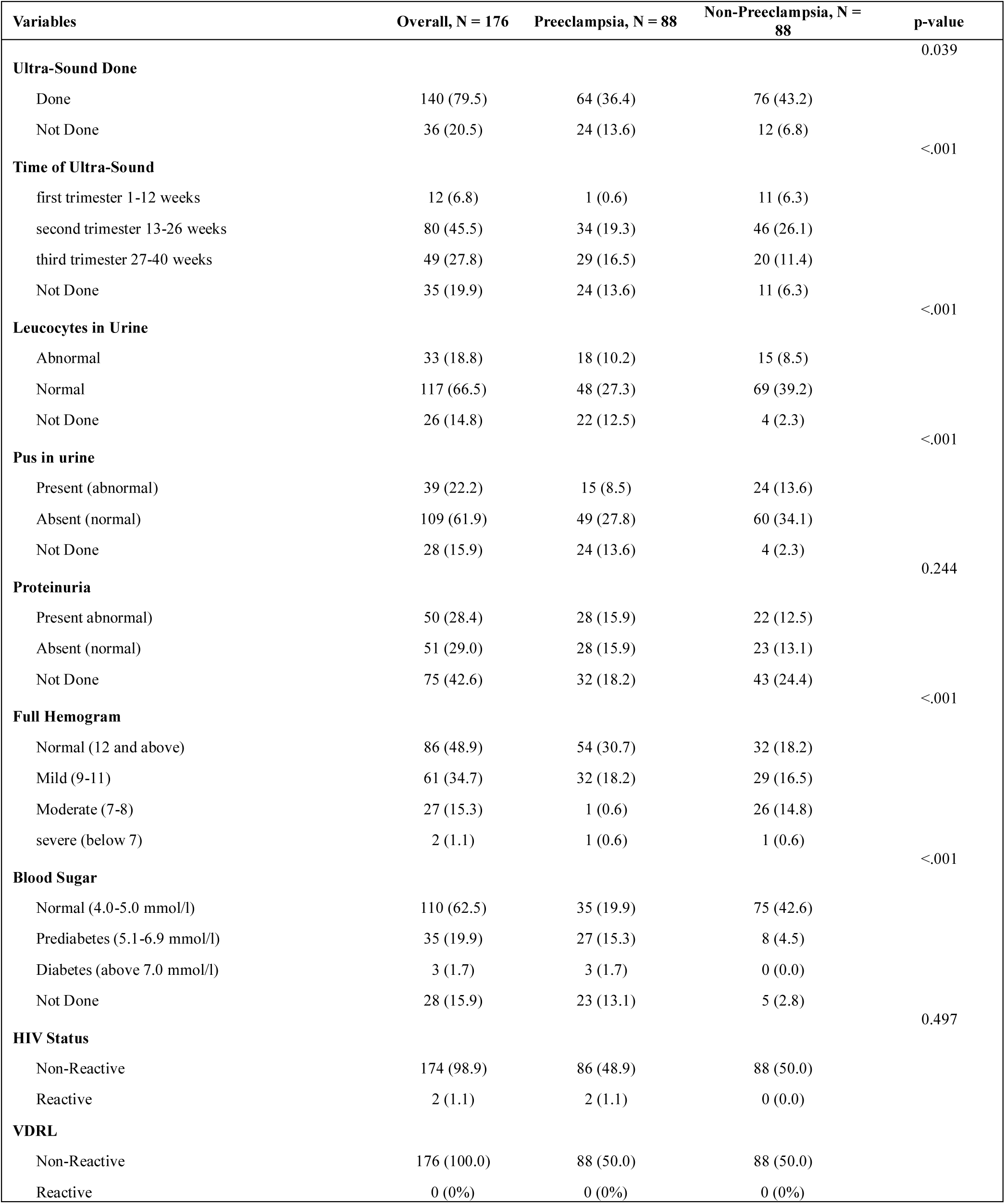
Paraclinical Tests Associated with Preeclampsia

| Variables | Overall, N = 176 | Preeclampsia, N = 88 | Non-Preeclampsia, N = 88 | p-value |
| --- | --- | --- | --- | --- |
| **Ultra-Sound Done** |  |  |  | 0.039 |
| Done | 140 (79.5) | 64 (36.4) | 76 (43.2) |  |
| Not Done | 36 (20.5) | 24 (13.6) | 12 (6.8) |  |
| **Time of Ultra-Sound** |  |  |  | <.001 |
| first trimester 1-12 weeks | 12 (6.8) | 1 (0.6) | 11 (6.3) |  |
| second trimester 13-26 weeks | 80 (45.5) | 34 (19.3) | 46 (26.1) |  |
| third trimester 27-40 weeks | 49 (27.8) | 29 (16.5) | 20 (11.4) |  |
| Not Done | 35 (19.9) | 24 (13.6) | 11 (6.3) |  |
| **Leucocytes in Urine** |  |  |  | <.001 |
| Abnormal | 33 (18.8) | 18 (10.2) | 15 (8.5) |  |
| Normal | 117 (66.5) | 48 (27.3) | 69 (39.2) |  |
| Not Done | 26 (14.8) | 22 (12.5) | 4 (2.3) |  |
| **Pus in urine** |  |  |  | <.001 |
| Present (abnormal) | 39 (22.2) | 15 (8.5) | 24 (13.6) |  |
| Absent (normal) | 109 (61.9) | 49 (27.8) | 60 (34.1) |  |
| Not Done | 28 (15.9) | 24 (13.6) | 4 (2.3) |  |
| **Proteinuria** |  |  |  | 0.244 |
| Present abnormal) | 50 (28.4) | 28 (15.9) | 22 (12.5) |  |
| Absent (normal) | 51 (29.0) | 28 (15.9) | 23 (13.1) |  |
| Not Done | 75 (42.6) | 32 (18.2) | 43 (24.4) |  |
| **Full Hemogram** |  |  |  | <.001 |
| Normal (12 and above) | 86 (48.9) | 54 (30.7) | 32 (18.2) |  |
| Mild (9-11) | 61 (34.7) | 32 (18.2) | 29 (16.5) |  |
| Moderate (7-8) | 27 (15.3) | 1 (0.6) | 26 (14.8) |  |
| severe (below 7) | 2 (1.1) | 1 (0.6) | 1 (0.6) |  |
| **Blood Sugar** |  |  |  | <.001 |
| Normal (4.0-5.0 mmol/l) | 110 (62.5) | 35 (19.9) | 75 (42.6) |  |
| Prediabetes (5.1-6.9 mmol/l) | 35 (19.9) | 27 (15.3) | 8 (4.5) |  |
| Diabetes (above 7.0 mmol/l) | 3 (1.7) | 3 (1.7) | 0 (0.0) |  |
| Not Done | 28 (15.9) | 23 (13.1) | 5 (2.8) |  |
| **HIV Status** |  |  |  | 0.497 |
| Non-Reactive | 174 (98.9) | 86 (48.9) | 88 (50.0) |  |
| Reactive | 2 (1.1) | 2 (1.1) | 0 (0.0) |  |
| **VDRL** |  |  |  |  |
| Non-Reactive | 176 (100.0) | 88 (50.0) | 88 (50.0) |  |
| Reactive | 0 (0%) | 0 (0%) | 0 (0%) |  |

The table 5 above showed that the type of delivery and baby sex did not differ significantly between groups (p=0.364 and p=0.880, respectively). Preeclampsia was significantly associated with lower birth weight (RR=1.97; OR=6.25, p<0.001), indicating substantially greater risk of low birth weight. APGAR scores also differed significantly (RR=1.85; OR=7.38, p=0.011), with poorer scores more frequent among babies of women with preeclampsia.

**Table 5:** Association Between Preeclampsia And Outcomes

| Variables | Overall, N = 176 | Preeclampsia, N = 88 | Non-Preeclampsia, N = 88 | p-value | Relative Risk/Odd Ratio |
| --- | --- | --- | --- | --- | --- |
| **Type of Delivery** |  |  |  | 0.364 |  |
| SVD | 95 (54.0) | 44 (25.0) | 51 (29.0) |  |  |
| C/S | 81 (46.0) | 44 (25.0) | 37 (21.0) |  |  |
| **Baby Weight** |  |  |  | 0.000 | 1.97 (6.25) |
| Exposed | 39 (22.2) | 31(17.6) | 8 (4.5) |  |  |
| Non-Exposed | 137 (77.8) | 57(32.4) | 80 (45.5) |  |  |
| **Baby Sex** |  |  |  | 0.880 |  |
| male | 94 (53.4) | 48 (27.3) | 46 (26.1) |  |  |
| female | 82 (46.6) | 40 (22.7) | 42 (23.9) |  |  |
| **APGAR Score** |  |  |  | 0.011 | 1.85 (7.38) |
| Poor | 15 (9.7) | 13 (7.4) | 4 (2.3) |  |  |
| Good | 158 (90.3) | 75 (42.6) | 84 (47.7) |  |  |

## Data Availability

All data included in this present study will be availed under reasonable request.

## Consent for Publication

The authors provided consent for publication

## Ethical Approval and Consent

Before enrollment, participants received information on their rights, voluntary participation, withdrawal, confidentiality, privacy, data management, and storage. Written informed consent and caregiver assent were obtained before inclusion.

## Availability of Data and Materials

Data are available after reasonable request

## Recommendations

Multicenter prospective cohort studies should be undertaken to establish causal pathways of preeclampsia in Kenyan populations.

## Authors Contributions

JM led the study conception, design, analysis, manuscript drafting, and supervision. DM supported study design, data analysis, and manuscript review. WM, BM, EW, PK, RN, JW contributed to implementation, data collection, and quality assurance. All authors approved and accepted responsibility for the final manuscript. JW supported the data entry/cleaning, statistical analysis, visualization, results interpretation, and manuscript review.

## Acknowledgement

The authors gratefully acknowledge the generous financial support provided by the Royal Society of Tropical Medicine and Hygiene (RSTMH), which made this study possible. The authors also sincerely appreciate the invaluable support of all study participants, healthcare workers, and Kiambu Counties level five management Teams.

## Abbreviations

NACOSTI: National Commission for Science, Technology and Innovation

APGAR: Appearance, Pulse, Grimace, Activity and Respiration

## References

1. Hutcheon JA, Lisonkova S, Joseph KS. Epidemiology of pre-eclampsia and the other hypertensive disorders of pregnancy. Best Pract Res Clin Obstet Gynaecol [Internet]. 2011 [cited 2026 Aug 20];25(4):391–403. Available from: https://pubmed.ncbi.nlm.nih.gov/21333604/

2. WHO. Pre-eclampsia [Internet]. 2025 [cited 2026 Aug 20]. Available from: https://www.who.int/news-room/fact-sheets/detail/pre-eclampsia

3. Vera-Ponce VJ, Loayza-Castro JA, Ballena-Caicedo J, Valladolid-Sandoval LAM, Zuzunaga-Montoya FE, Gutierrez De Carrillo CI. Global prevalence of preeclampsia, eclampsia, and HELLP syndrome: a systematic review and meta-analysis. Front Reprod Heal [Internet]. 2025 [cited 2026 Aug 26];7. Available from: https://pubmed.ncbi.nlm.nih.gov/41293035/

4. Atluri N, Beyuo TK, Oppong SA, Moyer CA, Lawrence ER. Challenges to diagnosing and managing preeclampsia in a low-resource setting: A qualitative study of obstetric provider perspectives from Ghana. PLOS Glob Public Heal [Internet]. 2023 May 1 [cited 2026 Aug 26];3(5):e0001790. Available from: https://journals.plos.org/globalpublichealth/article?id=10.1371/journal.pgph.0001790

5. Kenya National Bureau of Statistics (KNBS). Kenya Demographic and Health Survey 2022 [Internet]. 2022 [cited 2026 Aug 26]. Available from: https://www.knbs.or.ke/reports/kdhs-2022/

6. Ndwiga C, Odwe G, Pooja S, Ogutu O, Osoti A, Warren CE. Clinical presentation and outcomes of pre-eclampsia and eclampsia at a national hospital, Kenya: A retrospective cohort study. PLoS One [Internet]. 2020 Jun 1 [cited 2026 Aug 26];15(6):e0233323. Available from: https://journals.plos.org/plosone/article?id=10.1371/journal.pone.0233323

7. National Council for Population and Development. State of Kenya Population Report 2024. Nairobi, Kenya; 2024.

8. Ministry of Health. Kenya Maternal and Perinatal Death Surveillance and Response (MPDSR) Report. . Nairobi; 2025.

9. Kiambu County Government. County Integrated Development Plan 2023-2027 – Kiambu County Government [Internet]. 2023 [cited 2026 Aug 26]. Available from: https://kiambu.go.ke/downloads/county-integrated-development-plan-2023-2027/

10. Kim H-Y. Statistical notes for clinical researchers: Sample size calculation 2. Comparison of two independent proportions. Restor Dent Endod [Internet]. 2016 [cited 2026 Oct 5];41(2):154. Available from: https://pubmed.ncbi.nlm.nih.gov/27200285/

11. Duckitt K, Harrington D. Risk factors for pre-eclampsia at antenatal booking: systematic review of controlled studies. BMJ [Internet]. 2005 Mar 12 [cited 2026 Aug 26];330(7491):565–7. Available from: https://pubmed.ncbi.nlm.nih.gov/15743856/

12. Kiondo P, Wamuyu-Maina G, Bimenya GS, Tumwesigye NM, Wandabwa J, Okong P. Risk factors for pre-eclampsia in Mulago Hospital, Kampala, Uganda. Trop Med Int Health [Internet]. 2012 Apr [cited 2026 Aug 26];17(4):480–7. Available from: https://pubmed.ncbi.nlm.nih.gov/22151898/

13. Ayele AD, Tilahun ZA. Determinants of pre-eclampsia among women attending delivery services in public health institutions of Debre Tabor Town: a case-control study. Reprod Health [Internet]. 2022 Dec 1 [cited 2026 Aug 26];19(1). Available from: https://pubmed.ncbi.nlm.nih.gov/35804383/

14. Ezugwu EC, Mbah BO, Chigbu CO, Onah HE. Anaemia in pregnancy: a public health problem in Enugu, southeast Nigeria. J Obstet Gynaecol [Internet]. 2013 Jul [cited 2026 Aug 26];33(5):451–4. Available from: https://pubmed.ncbi.nlm.nih.gov/23815194/

15. Mrema D, Lie RT, Østbye T, Mahande MJ, Daltveit AK. The association between pre pregnancy body mass index and risk of preeclampsia: a registry based study from Tanzania. BMC Pregnancy Childbirth [Internet]. 2018 Feb 21 [cited 2026 Aug 27];18(1). Available from: https://pubmed.ncbi.nlm.nih.gov/29466949/

16. Hahka TM, Slotkowski RA, Akbar A, Vanormer MC, Sembajwe LF, Ssekandi AM, et al. Hypertension Related Co-Morbidities and Complications in Women of Sub-Saharan Africa: A Brief Review. Circ Res. 2024 Feb 16;134(4):459–73.

17. Phipps E, Prasanna D, Brima W, Jim B. Preeclampsia: Updates in Pathogenesis, Definitions, and Guidelines. Clin J Am Soc Nephrol [Internet]. 2016 Jun 6 [cited 2026 Aug 27];11(6):1102. Available from: https://pmc.ncbi.nlm.nih.gov/articles/PMC4891761/

18. Rana S, Lemoine E, Granger J, Karumanchi SA. Preeclampsia: Pathophysiology, Challenges, and Perspectives. Circ Res [Internet]. 2019 Mar 29 [cited 2026 Aug 27];124(7):1094–112. Available from: https://pubmed.ncbi.nlm.nih.gov/30920918/

19. Adjei S, Jnr WSA, Boamah RAB, Gyimah EO, Payne WK, Diawuo HY, et al. Pre- Eclampsia and Its Associated Factors Among Pregnant Women in Ghana: A Retrospective Study. Heal Sci Reports [Internet]. 2026 Aug 1 [cited 2026 Aug 27];9(8):e72990. Available from: https://pmc.ncbi.nlm.nih.gov/articles/PMC13442918/

20. Kokori E, Aderinto N, Olatunji G, Komolafe R, Babalola EA, Isarinade DT, et al. Prevalence and materno-fetal outcomes of preeclampsia/eclampsia among pregnant women in Nigeria: a systematic review and meta-analysis. Eur J Med Res [Internet]. 2024 Oct 3 [cited 2026 Aug 27];29(1):482. Available from: https://pmc.ncbi.nlm.nih.gov/articles/PMC11448017/

21. Ejaz S, Yaqoob R, Alkahtani R, Khalique AA, Azam M, Mahmood N, et al. The Association Of Socioeconomic Status With Preeclampsia In Pakistani Females. BioSight [Internet]. 2021 Dec 31 [cited 2026 Aug 27];2(2):50–7. Available from: https://biosight.org/index.php/bios/article/view/54

22. Turekulova A, Dzhardemaliyeva N, Mereke A, Kulimbet M. Maternal Disorders Associated with Morbidity and Mortality in a Metropolis of Kazakhstan. Clin Pract [Internet]. 2025 Jun 1 [cited 2026 Aug 27];15(6):108. Available from: https://pmc.ncbi.nlm.nih.gov/articles/PMC12191608/

23. World Health Organization (WHO). Target product profile for drugs to prevent pre-eclampsia [Internet]. 2023 [cited 2026 Aug 27]. Available from: https://www.who.int/publications/i/item/9789240081130

24. Demissie Beketie E, Tesfaye Tafese W, Zeleke Shiferaw B, Asfaw Tilahun G, Alemayehu Gebretsadik M, Desalegn Suraj K, et al. Determinants of preeclampsia among mothers attending perinatal care in Gurage zone public hospitals, Ethiopia, matched case control study. Int J Africa Nurs Sci [Internet]. 2022 Jan 1 [cited 2026 Aug 27];17:100453. Available from: https://www.sciencedirect.com/science/article/pii/S2214139122000609

25. Vigil-De Gracia P, Vargas C, Sánchez J, Collantes-Cubas J. Preeclampsia: Narrative review for clinical use. Heliyon [Internet]. 2023 Mar 1 [cited 2026 Aug 27];9(3):e14187. Available from: https://pmc.ncbi.nlm.nih.gov/articles/PMC10009735/

26. Gestational Hypertension and Preeclampsia: ACOG Practice Bulletin, Number 222. Obstet Gynecol [Internet]. 2020 Jun 1 [cited 2026 Aug 26];135(6):E237–60. Available from: https://pubmed.ncbi.nlm.nih.gov/32443079/

27. Tita AT, Szychowski JM, Boggess K, Dugoff L, Sibai B, Lawrence K, et al. Treatment for Mild Chronic Hypertension during Pregnancy. N Engl J Med [Internet]. 2022 May 12 [cited 2026 Aug 27];386(19):1781–92. Available from: https://www.nejm.org/doi/pdf/10.1056/NEJMoa2201295

28. Yang Y, Wu N. Gestational Diabetes Mellitus and Preeclampsia: Correlation and Influencing Factors. Front Cardiovasc Med [Internet]. 2022 Feb 16 [cited 2026 Aug 27];9:831297. Available from: https://pmc.ncbi.nlm.nih.gov/articles/PMC8889031/

29. International Diabetes Federation. Gestational Diabetes: Symptoms, Risks & Management Explained [Internet]. 2023 [cited 2026 Aug 27]. Available from: https://idf.org/about-diabetes/gestational-diabetes/

30. Lowe WL, Scholtens DM, Kuang A, Linder B, Lawrence JM, Lebenthal Y, et al. Hyperglycemia and adverse Pregnancy Outcome follow-up study (HAPO FUS): Maternal gestational diabetes mellitus and childhood glucose metabolism. Diabetes Care [Internet]. 2019 Mar 1 [cited 2026 Aug 27];42(3):372–80. Available from: https://pmc.ncbi.nlm.nih.gov/articles/PMC6385693/

31. Gilboa I, Kupferminc M, Schwartz A, Landsberg Ashereh Y, Yogev Y, Rappaport Skornik A, et al. The Association between Advanced Maternal Age and the Manifestations of Preeclampsia with Severe Features. J Clin Med 2023, Vol 12, Page 6545 [Internet]. 2023 Oct 16 [cited 2026 Aug 27];12(20):6545. Available from: https://www.mdpi.com/2077-0383/12/20/6545/htm

32. Poon LC, Shennan A, Hyett JA, Kapur A, Hadar E, Divakar H, et al. The International Federation of Gynecology and Obstetrics (FIGO) initiative on pre-eclampsia: A pragmatic guide for first-trimester screening and prevention. Int J Gynaecol Obstet [Internet]. 2019 May 1 [cited 2026 Aug 27];145 Suppl 1(Suppl 1):1–33. Available from: https://pubmed.ncbi.nlm.nih.gov/31111484/

33. Belay AS, Wudad T. Prevalence and associated factors of pre-eclampsia among pregnant women attending anti-natal care at Mettu Karl referal hospital, Ethiopia: cross-sectional study. Clin Hypertens [Internet]. 2019 Jul 1 [cited 2026 Aug 26];25(1). Available from: https://pubmed.ncbi.nlm.nih.gov/31304042/

34. Nathan HL, Seed PT, Hezelgrave NL, De Greeff A, Lawley E, Conti-Ramsden F, et al. Maternal and perinatal adverse outcomes in women with pre-eclampsia cared for at facility-level in South Africa: A prospective cohort study. J Glob Health [Internet]. 2018 [cited 2026 Aug 27];8(2):020401. Available from: https://pmc.ncbi.nlm.nih.gov/articles/PMC6076583/

35. Chege SW. Determinants of poor perinatal outcomes among babies born of mothers with preeclampsia at Mama Lucky Kibaki hospital, Nairobi county. MSc Adv Pract Midwifery [Internet]. 2023 Dec 1 [cited 2026 Aug 27]; Available from: https://ecommons.aku.edu/etd_ke_sonam_ms-apm/6

36. Morimoto K, O’Rourke L. Third Trimester Lower Extremity Lymphorrhea. Case Rep Obstet Gynecol [Internet]. 2021 [cited 2026 Aug 27];2021:3594923. Available from: https://pmc.ncbi.nlm.nih.gov/articles/PMC8645402/

37. WHO. WHO recommendations on maternal and newborn care for a positive postnatal experience. WHO Recomm Matern newborn care a Posit postnatal Exp. 2022;124–7.

38. Kumar A, Raj D, Gupta A, Kumar A. Assessment of knowledge of obstetric danger signs and its associated factors among pregnant women attending antenatal clinic of rural health training centre of a medical college: A cross-sectional study from Rajasthan. J Fam Med Prim Care. 2022 Oct;11(10):6487–92.

39. Akinola O, Zimba R, Banda K, Mangani CL, Shakwelele H, Abdallah I, et al. A systematic review of implementation strategies to improve timely initiation of antenatal care among pregnant women in sub-Saharan Africa. J Public Health Africa [Internet]. 2026 Mar 26 [cited 2026 Aug 27];17(1):1461. Available from: https://pmc.ncbi.nlm.nih.gov/articles/PMC13058560/

40. Foxman B. Urinary tract infection syndromes: occurrence, recurrence, bacteriology, risk factors, and disease burden. Infect Dis Clin North Am [Internet]. 2014 Mar [cited 2026 Aug 26];28(1):1–13. Available from: https://pubmed.ncbi.nlm.nih.gov/24484571/

41. Kalinderi K, Delkos D, Kalinderis M, Athanasiadis A, Kalogiannidis I. Urinary tract infection during pregnancy: current concepts on a common multifaceted problem. J Obstet Gynaecol [Internet]. 2018 May 19 [cited 2026 Aug 27];38(4):448–53. Available from: https://pubmed.ncbi.nlm.nih.gov/29402148/

42. Easter SR, Cantonwine DE, Zera CA, Lim KH, Parry SI, McElrath TF. Urinary tract infection during pregnancy, angiogenic factor profiles, and risk of preeclampsia. Am J Obstet Gynecol [Internet]. 2016 Mar 1 [cited 2026 Aug 27];214(3):387.e1-387.e7. Available from: https://www.sciencedirect.com/science/article/abs/pii/S0002937815012156

43. Buciu VB, Ciurescu S, Șerban DM, Novacescu D, Nicoleta N, Tomescu L, et al. The Compounded Risk of Maternal Anemia and Preeclampsia: Neonatal Outcomes and Predictive Modeling in a Low-Resource Tertiary Center. J Clin Med [Internet]. 2025 Jul 1 [cited 2026 Aug 27];14(14):5051. Available from: https://pmc.ncbi.nlm.nih.gov/articles/PMC12294900/

44. Committee ADAPP. 2. Diagnosis and Classification of Diabetes: Standards of Care in Diabetes—2024. Diabetes Care [Internet]. 2023 Jan 1 [cited 2026 Aug 26];47(Suppl 1):S20. Available from: https://pmc.ncbi.nlm.nih.gov/articles/PMC10725812/

45. Wu P, Green M, Myers JE. Hypertensive disorders of pregnancy. BMJ [Internet]. 2023 Jun 30 [cited 2026 Aug 27];381. Available from: https://www.bmj.com/content/381/bmj-2022-071653

46. Mwaniki MC, Omwodo KA, Ogutu O, Pulei A, Kosgei RJ, Nyakundi GG, et al. Comparing the risk of maternal and perinatal complications among women with hypertensive disorders of pregnancy to normotensive women: an institution-based cohort review in Kenya. BMC Pregnancy Childbirth [Internet]. 2025 Dec 1 [cited 2026 Aug 27];25(1):1166. Available from: https://pmc.ncbi.nlm.nih.gov/articles/PMC12587589/

47. Maharaj NR, Phulukdaree A, Nagiah S, Ramkaran P, Tiloke C, Chuturgoon AA. Pro-Inflammatory Cytokine Levels in HIV Infected and Uninfected Pregnant Women with and without Preeclampsia. PLoS One [Internet]. 2017 Jan 1 [cited 2026 Aug 27];12(1):e0170063. Available from: https://journals.plos.org/plosone/article?id=10.1371/journal.pone.0170063

48. Wimalasundera RC, Larbalestier N, Smith JH, De Ruiter A, Thom SAMG, Hughes AD, et al. Pre-eclampsia, antiretroviral therapy, and immune reconstitution. Lancet [Internet]. 2002 Oct 12 [cited 2026 Aug 27];360(9340):1152–4. Available from: https://pubmed.ncbi.nlm.nih.gov/12387967/

49. Suy A, Martínez E, Coll O, Lonca M, Palacio M, De Lazzari E, et al. Increased risk of pre-eclampsia and fetal death in HIV-infected pregnant women receiving highly active antiretroviral therapy. AIDS. 2006;20(1):59–66.

50. The International Federation of Gynecology and Obstetrics (FIGO). FIGO releases new guidelines to combat Pre-Eclampsia | Figo [Internet]. 2019 [cited 2026 Aug 27]. Available from: https://www.figo.org/figo-releases-new-guidelines-combat-pre-eclampsia

51. Duncan JR, Odibo AO. Controversies in the management of fetal growth restriction: a state-of-the-art review. J Matern Neonatal Med [Internet]. 2026 Dec 31 [cited 2026 Aug 27];39(1):2717471. Available from: https://www.tandfonline.com/doi/pdf/10.1080/14767058.2026.2717471

52. Osoro E, Awuor AO, Inwani I, Mugo C, Hunsperger E, Verani JR, et al. Association between low maternal serum aflatoxin B1 exposure and adverse pregnancy outcomes in Mombasa, Kenya, 2017–2019: A nested matched case–control study. Matern Child Nutr. 2024 Oct 1;20(4).

53. National Institute for Health and Care Excellence (NICE). Overview | Hypertension in pregnancy: diagnosis and management | Guidance | NICE. 2023;

54. Koopmans CM, Bijlenga D, Groen H, Vijgen SM, Aarnoudse JG, Bekedam DJ, et al. Induction of labour versus expectant monitoring for gestational hypertension or mild pre-eclampsia after 36 weeks’ gestation (HYPITAT): a multicentre, open-label randomised controlled trial. Lancet [Internet]. 2009 Sep 25 [cited 2026 Aug 27];374(9694):979–88. Available from: https://pubmed.ncbi.nlm.nih.gov/19656558/

55. Lawn JE, Ohuma EO, Bradley E, Okwaraji YB, Yargawa J, Blencowe H, et al. Small babies, big risks: global estimates of prevalence and mortality for vulnerable newborns to accelerate change and improve counting. Lancet (London, England) [Internet]. 2023 May 20 [cited 2026 Aug 27];401(10389):1707–19. Available from: https://pubmed.ncbi.nlm.nih.gov/37167989/

56. Chawanpaiboon S, Titapant V, Pooliam J. Neonatal complications and risk factors associated with assisted vaginal delivery. Sci Rep [Internet]. 2024 Dec 1 [cited 2026 Aug 27];14(1):11960. Available from: https://pmc.ncbi.nlm.nih.gov/articles/PMC11127920/

